# Prevalence of Healthy Eating Initiatives in Australian Primary Schools: A Cross-Sectional Survey

**DOI:** 10.1101/2025.10.27.25338934

**Authors:** Jessica Bell, Kate M O’Brien, Luke Wolfenden, Serene Yoong, Adrian Bauman, Lucy Leigh, Christophe Lecathelinais, Rebecca K Hodder

## Abstract

**Introduction:** Schools are recommended settings for promotion of healthy eating, but to achieve their intended impact, initiatives must be implemented population wide. There is currently limited data available to assess the implementation of Australian school-based healthy eating initiatives.

**Objective:** To assess the implementation of healthy eating initiatives in Australian primary schools and whether implementation is associated with school characteristics.

**Methods:** A cross-sectional study of a nationally representative sample of Australian primary school principals from August 2022 to October 2023. Principals reported on implementation of 32 healthy eating initiatives grouped by guideline-informed opportunities: healthy food in the classroom, at school, brought to school, and outside of school. Prevalence estimates were weighted using iterative ranking to reflect population characteristics. Logistic regression models assessed the associations between initiatives and school characteristics.

**Results:** Principals or their representatives from 569 Australian schools completed the survey. Prevalence of initiative implementation varied considerably, ranging from 7% to 97%. The least and most frequently implemented initiative in the classroom was ‘Teachers acted as healthy eating role models for students’ and ‘Students supported to be healthy eating role models for peers’ (both 8%) and ‘*Permission or breaks to drink water in class time in ≥80% of classes daily*’ (97%); to increase healthy food at school was ‘*Provide incentives or rewards to children for purchasing/choosing healthy foods or drinks at school’* (11%) and ‘*Install water stations for free access to cooled plain or optionally carbonated water’ (64%)*; to improve healthy food brought to school was ‘*Workshops with parents on healthy eating’* (14%) and ‘*Information on healthy eating sent home to parents*’ (88%); and to improve healthy eating outside of school was ‘*Promote healthy food options at shops close to school*’ and ‘*Community members acted as healthy eating role models for students’* (both 15%) and ‘*Healthy eating programs delivered in partnership with community organisations or services*’ (31%). A range of initiatives were associated with school size (5 initiatives) including ‘*Provide free fruits and vegetables to children’ and* ‘*Provide free healthy school lunches*’; geographic remoteness (7 initiatives) such as ‘*Teachers acted as healthy eating role models for students*’ and ‘Imple*ment healthy cooking programs or similar healthy eating experiences’*, or socio-economic status of school locality (3 initiatives) such as *‘Implement healthy fundraising policies’ and* ‘*Provide free breakfast to children’*.

**Conclusion:** This first national study to comprehensively assess the implementation levels of a broad range of healthy eating initiatives reveals key insights into where policy and practice support is needed. Implementation of the initiatives varied significantly, with some associations with school characteristics. Initiatives with low implementation identifies opportunities for further investigation.

## 1. Introduction

Healthy eating is recognised as a key determinant for preventing obesity, and non-communicable diseases more broadly.^1^ A healthy diet, which emphasises the consumption of fruits, vegetables, whole grains, and lean protein, is integral to maintaining healthy and wellbeing.^2^ Influencing dietary habits early in life is particularly important as many healthy eating behaviours are established during childhood and track into adulthood.^3^ As a result, establishing a healthy diet during childhood is identified as a priority to mitigate the burden of obesity.^4–6^ Guidelines for healthy eating, such as those produced by the World Health Organization (WHO) and Australian Department of Health, Disability and Aging provide evidence-based recommendations regarding optimal dietary intake for children.^2, 7^ However, adherence to this guidance is generally poor.^5, 8^ In Australia, only 9% of children in 2020/21 aged 2-17 years met the recommendation for servings of fruits and vegetables.^9^

Schools play a crucial role in shaping children’s diet and eating behaviours, as they provide an environment that can support or hinder healthy choices. As such, schools are widely recommended as key settings for the promotion of healthy eating due to their infrastructure and extensive reach.^10–12^ In line with this, there has been significant investment in identifying effective school-based healthy eating initiatives. School-based healthy eating initiatives have been found to be effective by many high-quality systematic and umbrella reviews in improving dietary outcomes.^13–16^ For example, school-based healthy eating initiatives such as nutrition education, school food environment policies and fruit and vegetable provision have been shown to improve intake of fruit and vegetables and reduce the consumption of sugar-sweetened beverages.^13, 15, 16^

As child nutrition behaviours are impacted by a range of social and environmental factors, healthy eating initiatives in schools are typically multicomponent, and include combinations of initiatives targeting nutritional education, healthier food policies, and supportive environments to promote healthy eating behaviours.^17^ To support schools, Australian governments and health organisations have developed a range of policies, toolkits and programs promoting nutritious food environments. For example, the *Good Practice Guide* from the Council of Australian Governments outlines national principles for supporting healthy eating and drinking in schools through a whole-of-school approach.^18^ Several states and territories have also developed their own resources and guidelines. In New South Wales, the *Live Life Well @ School* program promotes curriculum-linked nutrition education, supportive environments, and partnerships with families.^19^ Similarly, Queensland’s *Healthy Start to School Toolkit*,^20^ Western Australia’s *Healthy Schools Program Toolkit* ^21^ and Health Promoting Schools Toolkit,^21^ and Victoria’s Healthy Eating Advisory Service resources^22^ provide strategies to integrate healthy eating into school policies, curriculum, and the physical and social environment. For the intended public health impact of effective school-based initiatives to be achieved however, they need to be implemented at scale.

There is currently no national data on the healthy eating initiatives that are implemented in Australian primary schools, however, there is some state-level data available. For example, between 2006 and 2013, Nathan et al. examined healthy eating initiatives in NSW primary schools, including a longitudinal study of four initiatives across 303 schools. In 2013, the study reported 39% of schools had a healthy eating policy, 73% integrated teaching of healthy eating education across other subjects, 89% offered fruit and vegetable breaks, and 96% promoted healthy eating to families.^23, 24^ Similar levels of implementation of healthy initiatives embedded within the *Live Life Well @ School* program in a sample of 1707 NSW primary schools in 2015 (e.g. 84% of schools implemented fruit and vegetable breaks).^25^ In Queensland, a 2012 evaluation of the *Smart Choices* program found high implementation levels for school breakfast programs (98%), healthy events other than sporting (87%), with healthy fundraising, curriculum activities, and other events implemented by 80%, 97%, and 75% of schools. However, this state level data is limited as existing studies were typically undertaken prior to COVID-19, and most focused on a limited number of initiatives.

School food environments and healthy eating initiatives may vary based on a range of school characteristics, including location, size and socioeconomic status.^26^ For example, the 2015 NSW SPANS Report found that rural schools were more likely than urban schools to comply with healthy canteen policies (88% vs. 39%) and implement initiatives like fruit and vegetable break program *Crunch&Sip®*.^27^ Similarly, implementation levels of fruit and vegetable breaks as part of the NSW *Live Life Well @ School* program have been reported to differ for schools located in socio-economically disadvantaged or remote areas.^25^ However, to the authors’ knowledge there is no national data that reports implementation of school-based healthy eating initiatives by school characteristics.

To guide national strategies to improve public health nutrition there is a need for contemporary and comprehensive data regarding implementation of school-based healthy eating initiatives. Despite state-level insights, national data to comprehensively evaluate implementation across diverse school contexts remains lacking. To address these evidence gaps, a national cross-sectional study was conducted to determine the current implementation of a broad range of healthy eating initiatives in Australian primary schools and determine whether implementation is associated with school characteristics.

## 2. Methods

### 2.1 Study design

A cross-sectional study of Australian primary school principals was conducted between August 2022 and October 2023. The study received ethics approval from the University of Newcastle Human Research Ethics Committee (approval no. H-2021-0045) and research approval from 25 out of 32 Australian state and catholic school jurisdictions (Appendix 1).

### 2.2 Sample and recruitment

Australian schools with primary aged student enrolments from all jurisdictions were eligible for participation, and a nationally representative sample was selected. Non-mainstream schools (e.g. those for special purposes such as students with intellectual disabilities), language or mature age schools, dedicated preschools, schools providing short-term education, or schools within jurisdictions without research approval were not eligible.

The Australian Council for Education Research (ACER) selected a nationally representative random sample of 4500 Australian primary schools using equal-probability stratified sampling. Schools were stratified according to state/territory, sector (government, catholic, and independent schools), remoteness (very remote, remote, outer regional, inner regional, major cities), socio-economic status (using Socio-Economic Indexes for Areas (SEIFA) Index of Education (IEO) National Decile) and school size (total enrolment quartiles).

Principals of sampled schools or their nominated delegate were invited to participate in the study. Study information, which included an invitation and an online consent form was sent to a publicly-available school email address. The invitation allowed participants to nominate their preference complete the survey online (via the Research Electronic Data Capture platform (REDCap)^28^ or via a Computer Assisted Telephone Interview (CATI). Principals who opted to complete the survey online were sent a unique REDCap survey link. If CATI was preferred, participants were telephoned by a trained interviewer who entered responses into the REDCap platform. Both methods took approximately 30 minutes to complete.

An email prompt was sent to schools that had not completed the survey one week following invitation. One week later, schools that had not completed received up to 10 follow-up telephone prompts to complete the survey by trained interviewers before being considered as a non-response. Telephone prompts were reduced or omitted for three school jurisdictions. All eligible schools were invited to participate, and prompting of schools continued until the target quota for each state or territory was achieved.

### 2.3 Data collection and measures

#### 2.3.1 School characteristics

School characteristics were sourced from the ACER database of all Australian schools, and supplemented by data from the publicly available Australian Curriculum, Assessment and Reporting Authority^29^ and included: state/territory; education sector; jurisdiction or Archdiocese/Diocese; Australian Statistical Geography Standard (ASGS) remoteness category; socio-economic status; school size (total enrolments) and proportion who are Aboriginal and/or Torres Strait Islander.

#### 2.3.2 Participant characteristics

Participants reported their current position and length of time in the role.

#### 2.3.3 Implementation of healthy eating initiatives

Healthy eating initiatives were identified from recent high-quality systematic reviews of school-based healthy eating programs conducted globally and were selected to align with recommendations from Australian and international policies and guidelines.^30^ Participants were randomised (using stratified randomisation in SAS Version 9.4^31^ by state/territory and sector) to answer survey items regarding the healthy eating initiatives implemented within their school. To reduce participant burden, and in consideration of the large number of possible healthy eating initiatives being examined, schools were further randomised to survey arm 1 or 2 using stratification randomisation by state/territory and sector in SAS Version 9.4.^31^ This was conducted to ensure an even distribution of participants across the two arms.

Survey items asked principals to report whether their school was currently implementing 32 individual healthy eating initiatives (Appendix 2: Healthy eating initiative definitions). The healthy eating initiatives were grouped into five categories based on guideline recommended opportunities for healthy eating initiatives: ‘Healthy eating in the classroom’ (11 initiatives), ‘Healthy food available at school’ (11 initiatives) ‘Healthy food brought to school’ (3 initiatives) ‘Healthy eating outside the school or involving families’ (4 initiatives), and ‘Other’ (3 initiatives). Existing validated survey items^32^ were mapped to each healthy eating initiative, or where an existing validated item could not be identified, the research team with expertise in school-based implementation developed a new item to assess implementation (Appendix 3: Principal survey). The principal survey was piloted with research staff with teaching experience, stakeholders within the sector, and school principals following ethics approval. Modifications to question wording were made prior to survey commencement to improve interpretation and data quality.

### 2.4 Sample size

A sample of 350 schools was estimated to be sufficient to generate a weighted national prevalence of healthy eating initiative implementation with a precision of ±5.2%.

### 2.5 Statistical analysis

School postcodes were used to classify schools using the national SEIFA 2016 index of relative socio-economic disadvantage which focuses on relative socio-economic disadvantage.^33^ Descriptive statistics summarised school characteristics, staff demographic characteristics and participation rates. Chi-squared analyses were used to compare characteristics between participating and non-participating schools (state/territory, school type, remoteness, socio-economic status, school size).

The prevalence estimates of implementation of each healthy eating initiative were adjusted by raking the survey weights iteratively to align sample distributions with the Australian school population (taking into account the stratification variables jurisdiction, sector, SEIFA, Australian Statistical Geography Standard remoteness category, and school size quartile) and to mitigate non-response bias using SAS Version 9.4.^31^ Integrated and tailored weight trimming during raking iterations was applied, reducing excessive weights while maintaining alignment with population controls to generate the weight prevalence estimates for each healthy eating initiative.

Weighted binary logistic regression analyses were conducted to examine the univariate associations between the level of implementation of healthy eating initiatives and school characteristics: school size (small <300 / large ≥300), remoteness (urban: major city / rural: inner regional, outer regional, remote according to the Australian Statistical Geography Standard remoteness categories^34^) and socio-economic status (most disadvantaged / least disadvantaged according to the SEIFA Index of relative socio-economic disadvantage categories^33^). Odds ratios (OR) and 95% confidence levels (CI) were calculated for each model. Statistical analyses were performed using SAS Version 9.4.^31^ Statistical significance was set at an alpha level of 0.05.

## 3. Results

### 3.1 School and principal characteristics

There were 7793 Australian schools with primary school enrolments. Out of these, 514 were deemed ineligible (e.g. non-mainstream schools) prior to sampling. Of the 4500 schools identified via the stratified random sampling method, 1064 (24%) schools were classified as out of scope (see Appendix 4 for reasons these schools were considered out of scope), leaving 3436 eligible schools in the sampling frame. A total of 3303 primary schools were invited to participate in the survey, which was conducted between 9 August 2022 and 20 October 2023 (133 schools were not invited due to meeting quota for that state). In total, 669 schools completed the survey (via telephone n=57, online n=612; 20% participation rate) and 460 (14%) declined.

Of the 669, 569 schools completed at least one healthy eating survey item and the sample for each survey item ranged from 164 to 301. The characteristics of the schools and principals are reported in Table 1. Most participating schools were from the government sector (64%), located outside urban areas (55%), were situated in the most disadvantaged areas (54%), and were small schools (<300 students; 63%). The average student enrolment across the schools was 287 (range 2-1,703) students and 18% were Indigenous enrolments. Surveys were primarily completed by school principals (87%) and 55% of the schools had an operational canteen service. The characteristics of participating schools closely mirrored the overall distribution of primary schools in Australia, except for schools from NSW (Table 1). Additionally, a higher proportion of small schools from rural areas participated in the survey compared to their representation in the national primary school demographic. There were significant differences in the school characteristics of participating versus non-participating eligible schools for state (p<0.001), sector (p<0.001), remoteness (p<0.001), and school size (p<0.001).

**Table 1.** Schools and principal characteristics^a^.

| Characteristics | Participating schools N=569<br>n (%) | Invited schools N=4159 | Eligible Australian schools<sup>b</sup><br>(%) |
| --- | --- | --- | --- |
| **State/territory** |  |  |  |
| Australian Capital Territory | 10 (1.76) | 63 (1.51) | 1.39 |
| New South Wales | 58 (10.19) | 1313 (31.57) | 31.79 |
| Northern Territory | 13 (2.28) | 66 (1.59) | 2.16 |
| Queensland | 106 (18.63) | 561 (13.49) | 18.60 |
| South Australia | 64 (11.25) | 369 (8.87) | 7.88 |
| Tasmania | 24 (4.22) | 128 (3.08) | 2.77 |
| Victoria | 199 (34.97) | 1120 (26.93) | 23.72 |
| Western Australia | 95 (16.70) | 539 (12.96) | 11.69 |
| **Sector** |  |  |  |
| Government | 366 (64.32) | 3034 (72.95) | 69.29 |
| Catholic | 128 (22.50) | 616 (14.81) | 17.85 |
| Independent | 75 (13.18) | 509 (12.24) | 12.85 |
| **Remoteness** |  |  |  |
| Urban<sup>c</sup> | 256 (44.99) | 2154 (51.79) | 51.35 |
| Rural<sup>d</sup> | 313 (55.01) | 2005 (48.21) | 48.65 |
| **SEIFA National (IRSD)** |  |  |  |
| Most Disadvantaged | 309 (54.31) | 2193 (52.73) | 52.83 |
| Least Disadvantaged | 260 (45.69) | 1966 (47.27) | 47.17 |
| **School size** |  |  |  |
| Small (<300 students) | 356 (62.57) | 2228 (53.57) | 52.67 |
| Large (≥300 students) | 213 (37.43) | 1931 (46.43) | 47.33 |
| **Total enrolment, mean (SD), range** | 287.42 (275.06),<br>2 to 1703 | 342.05 (325.43),<br>2 to 3567 | 366.21 (380.78),<br>2 to 4726 |
| **Indigenous Enrolments, mean % (SD)** | 9.73 (18.34) | 10.22 (17.89) | 10.67 (18.55) |
| **Staff Role who completed the survey** |  |  |  |
| Principal<sup>e</sup> | 494 (86.82) | - | - |
| Deputy/Assistant principal<sup>f</sup> | 34 (5.98) | - | - |
| Head of school / Campus / Operations | 11 (1.93) | - | - |
| Lead / classroom teacher / Health and Wellbeing Teacher / Coordinator | 20 (3.51) | - | - |
| Other role<sup>g</sup> | 10 (1.76) | - | - |
| **Schools with operational canteen, mean (SD)** | 314 (55.18) | - | - |
| **How long been in this role** |  |  |  |
| 12 months or more, range in years | 448 (78.73), 1 to 40 | - | - |
| Less than 12 months | 121 (21.27) | - | - |

<sup>a</sup>Reported as n (%), unless otherwise stated
<sup>b</sup>Data from the publicly available Australian Curriculum, Assessment and Reporting Authority
<sup>c</sup>Includes major cities
<sup>d</sup>Includes inner regional, outer regional, remote and very remote
<sup>e</sup>Executive principal, principal, acting principal
<sup>f</sup>Lead teacher, classroom teacher, health and wellbeing teacher, coordinator
<sup>g</sup>Administration officer, business manager, school counsellor/social worker, school nurse
Abbreviations: ACT: Australian Capital Territory; QLD: Queensland; NSW: New South Wales; NT: Northern Territory; SA: South Australia; SD: standard deviation; SES: Socio-Economic Status; TAS: Tasmania; VIC: Victoria; WA: Western Australia

### 3.2 Weighted prevalence of implementation of healthy eating initiatives

The weighted prevalence of implemented healthy eating initiatives is described in Table 2. Of the 32 healthy eating initiatives, the weighted prevalence ranged from 7% for ‘*The majority of teaching staff participated in healthy eating professional development*’ to 97% for ‘*Permission or breaks to drink water in class time in ≥80% of classes daily*’.

**Table 2.**
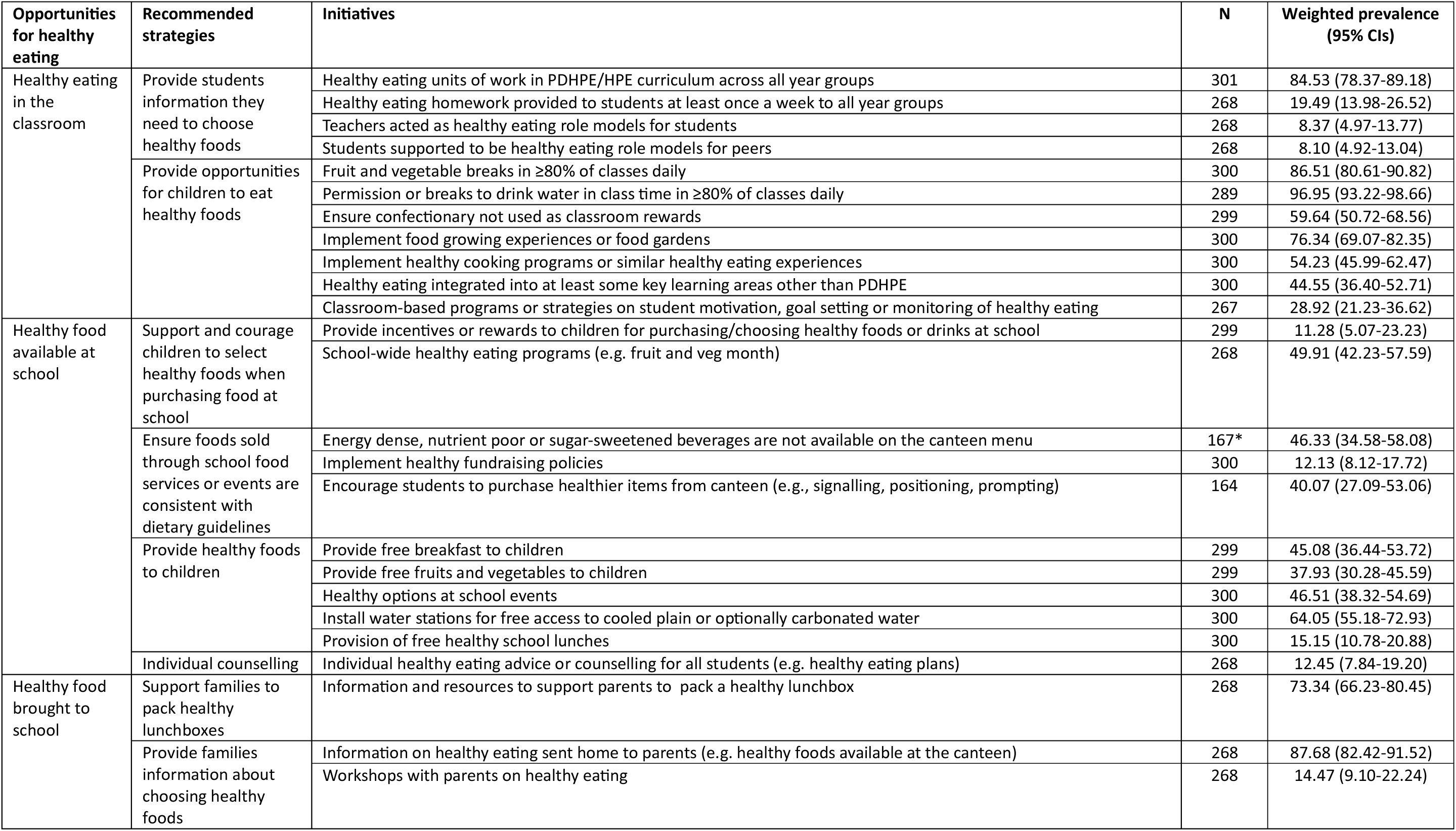
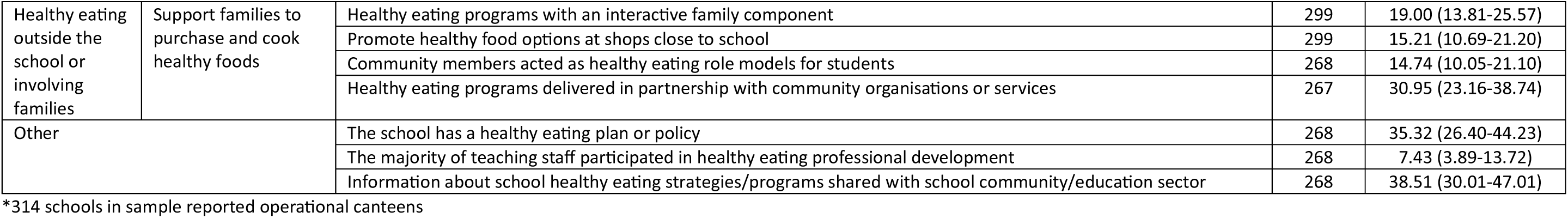
Weighted prevalence of implementation of healthy eating initiatives.

#### 3.2.1 Healthy eating in the classroom

For the 11 healthy eating initiatives implemented in the classroom, the weighted prevalence ranged from 8% (‘*Students supported to be healthy eating role models for peers’* and *‘Teachers acted as healthy eating role models for students’*) to 97% (‘*Permission or breaks to drink water in class time in ≥80% of classes daily’*). Other initiatives that were implemented in the majority of schools included ‘*Fruit and vegetable breaks in ≥80% of classes daily*’ (87%), *‘Healthy eating units of work in PDHPE/HPE curriculum across all year groups’* (85%), ‘*Implement food growing experiences or food gardens*’ (76%), ‘*Ensure confectionary not used as classroom rewards*’ (60%), and ‘*Implement healthy cooking programs or similar healthy eating experiences’* (54%).

#### 3.2.2 Healthy food available at school

The implementation of the 11 healthy eating initiatives categorised as ‘Healthy food available at school’ ranged from 11% (‘*Provide incentives or rewards to children for purchasing/choosing healthy foods or drinks at school*’) to 64% (‘*Install water stations for free access to cooled plain or optionally carbonated water*’). *‘School-wide healthy eating programs (e.g. fruit and veg month)’* (50%) was also implemented in half of the schools.

#### 3.2.3 Healthy food brought to school

The weighted prevalence of three initiatives to improve healthy food brought to school ranged from 14% (‘*Workshops with parents on healthy eating*’) to 88% (‘*Information on healthy eating sent home to parents (e.g. healthy foods available at the canteen)*’. ‘*Information and resources to support parents to pack a healthy lunchbox*’ was implemented in 73% schools.

#### 3.2.4 Healthy eating outside the school or involving families

Of the four healthy eating initiatives targeting improvements in healthy eating outside the school or involving families, none were implemented in ≥50% of schools: ‘*Healthy eating programs delivered in partnership with community organisations or services*’ (31%), ‘*Healthy eating programs with an interactive family component’* (19%), and ‘*Promote healthy food options at shops close to school’* and ‘*Community members acted as healthy eating role models for students*’ (both 15%).

#### 3.2.5 Other

Of the three healthy eating initiatives categorised as other, none were implemented in over 50% of schools: ‘*Information about school healthy eating strategies/programs shared with school community/education sector’* (35%), *‘The school has a healthy eating plan or policy’* (35%) and ‘The *majority of teaching staff participated in healthy eating professional development’* (7%).

### 3.3 Association between school characteristics and implementation of healthy eating initiatives

Associations between school size, remoteness or socio-economic status were found for 10 of the 32 initiatives summarised below (Appendix 5). Specifically, significant associations were found for five initiatives related to school size, seven initiatives related to remoteness and three initiatives related to socio-economic status.

#### 3.3.1 School size

Compared to large schools (≥300 enrolments), small schools had higher odds of implementing five initiatives. Small schools had higher odds of implementing ‘*Food growing experiences or food gardens’* (83% vs. 69%; OR 2.21 (95%CI 1.05–4.65)), *‘Provide free fruits and vegetables to children’* (46% vs. 28%; OR 2.19 (95%CI 1.14–4.21)), *‘Provision of free healthy school lunches’* (22% vs. 7%; OR 3.85 (95%CI 1.61–9.22)), *‘Healthy eating programs with an interactive family component’* (25% vs. 12%; OR 2.52 (95%CI 1.14–5.61)), and *‘Healthy eating programs delivered in partnership with community organisations or services’* (40% vs. 21%; OR 2.49 (95%CI 1.17–5.29)).

#### 3.3.2 Remoteness

Compared to schools in urban areas, schools in rural areas had higher odds of implementing six initiatives. Schools in rural areas had higher odds of implementing ‘*Healthy eating integrated into at least some key learning areas other than PDHPE’* (55% vs. 34%; OR 2.38 (95%CI 1.26–4.49)), *‘Provide free breakfast to children’* (55% vs. 35%; OR 2.29 (95%CI 1.06–4.92)), *‘Healthy eating programs with an interactive family component’* (25% vs. 13%; OR 2.15 (95%CI 1.02–4.53)), *‘Provision of free healthy school lunches’* (23% vs. 8%; OR 3.55 (95%CI 1.58–7.98)), *‘Implement healthy cooking programs or similar healthy eating experiences’* (65% vs. 44%; OR 2.23 (95%CI 1.16–4.74)), and *‘Provide free fruits and vegetables to children’* (51% vs. 26%; OR 2.98 (95%CI 1.55–5.71)). Compared to schools in rural areas, schools in urban areas had higher odds of implementing ‘*Teachers acted as healthy eating role models for students’* (14% vs. 2%; OR 7.14 (95%CI 2.17–25.00)).

#### 3.3.3 Socioeconomic status

Compared to schools in the least disadvantaged areas, schools in the most disadvantaged areas had higher odds of implementing two initiatives. Schools in the most disadvantaged areas had higher odds of implementing ‘*Provide free breakfast to children’* (60% vs. 30%; OR 3.70 (95%CI 1.85–7.14)) and*‘Healthy eating programs delivered in partnership with community organisations or services’* (38% vs. 22%; OR 2.22 (95%CI 1.06–4.76)). Compared to schools in the most disadvantaged areas, schools in the least disadvantaged areas had higher odds of implementing ‘*Healthy fundraising policies’* (18% vs. 6%; OR 3.26 (95%CI 1.28–8.29)).

## 4. Discussion

This study provides the first nationally representative snapshot of the prevalence of a broad range of evidence-based healthy eating initiatives in Australian primary schools, and their association with school characteristics. Implementation rates varied considerably, from 7% to 97% across the initiatives assessed. Of the 32 interventions assessed, nine were implemented in greater than 50% of schools, and five were implemented in over 75%. Implementation of 10 of the 32 initiatives were significantly associated with at least one school characteristic.

The implementation of healthy eating initiatives in Australian primary schools varied across the five categories. Of the nine initiatives implemented in greater than 50% of schools, six were delivered in classrooms. Classroom-based initiatives may be implemented in a greater proportion of primary schools due to alignment with existing teaching structures and curriculum requirements. Initiatives in the categories ‘*Healthy foods available at school*’, ‘*Healthy eating outside of school or involving family*’ or ‘*Other’* generally had lower levels of implementation. Evaluation of Australian healthy food and drink initiatives shows consistently higher uptake for classroom and school-based initiatives when compared to those which rely on family engagement, community partnerships, or broader systemic actions. For example, in the Western Australian Healthy Food & Drink School Principal Survey 2021, canteen menu compliance (removal of “red” items and meeting “green” targets) was reported by 89– 96% of metropolitan primary schools, but much lower levels were seen for initiatives involving families or outside-school contexts (only 33% of schools reported “sending healthy eating information home to families” and just 28% reported “hosting events promoting healthy eating”).^35^ These patterns of implementation may reflect multiple factors. For example, initiatives more easily influenced by the school, such as canteen menu changes, may be more amenable to implementation as the school directly manages resources, staff, and the environment. In contrast, initiatives delivered outside the school or involving families—such as workshops, home-based nutrition guidance, or community partnerships—face added complexity, relying on external actors, and logistical challenges. Further investigation of barriers to implementation of effective initiatives in these categories may be warranted to improve scaled implementation.

There is limited data on the implementation of healthy eating initiatives in Australian primary schools with which to make comparisons to the findings of our study. The results of this study were consistently lower than the available state-level data on the implementation of heathy eating initiatives compared to this study, which is representative of all Australian schools.^23, 27, 35, 36^ The 2013 study by Nathan et al. reported that 73% of schools implemented ‘integration of healthy eating into other subjects’ and that 89% of schools provided a ‘fruit and vegetable break’. In comparison, our study found that only 45% of schools embedded healthy eating into other subject areas and 87% of schools offered a fruit and vegetable break to students. Surveys conducted by state-based government organisations, such as the 2015 NSW SPANS survey also reported higher levels of implementation for the initiatives, including healthy eating in their curriculum (97% vs 85%) and communicating the importance of healthy eating to parents (94% vs 88%).^27^ The study conducted in 2021 by the Western Australia Department of Health reported that 79% of schools do not sell unhealthy foods at the canteens, even on an occasional basis, compared to the 46% of school canteens reported in this study. The findings of this study provide a comprehensive and contemporary assessment of the implementation of school-based healthy eating initiatives in Australian primary school and provides important data to guide future policy development.

In this study, significant associations were identified between school characteristics and the implementation of 18 initiatives. The findings reveal critical insights into the practice implementation by school size, remoteness, socioeconomic status. Smaller schools also consistently were found to have increased odds of implementation. Such findings may reflect the greater adaptability, autonomy and stronger community engagement of smaller schools to implement programs.^37^ Higher levels of implementation of obesity prevention initiatives among smaller schools has also been reported in other studies examining the implementation. Schools in rural areas also generally reported greater odds of implementation of nutrition initiatives compared to those in major cities and at times, so did schools in the most socio-economically disadvantaged areas. Such findings were surprising, given evidence of a geographic and socioeconomic gradient of healthy eating among Australian children. The findings may be due to reflect efforts by, or support to such schools to address these existing inequities in public health nutrition. Overall, these findings indicate the need for policymakers and program designers to consider contextual factors such as location, socioeconomic context, and school size when prioritising, designing, and implementing school-based initiatives.

A limitation of the study was the low response rate at just above 20%. Several strategies were employed to mitigate this limitation, including randomised sampling stratified according to jurisdiction and sector, with quotas set across each stratification. Statistical weighting according to the demographic profile of Australian primary schools was applied to the results to enhance representativeness. The anticipated quota for NSW government schools was not met, with only one of the estimated 161 schools participating in the survey, due to limitations in the research approval for this jurisdiction. As a result, the post-stratification weighting in NSW primarily relied on implementation data from Catholic and independent schools to calculate weighted implementation levels. It is unknown what impact this may have had on the estimates of weighted implementation across the healthy eating initiatives, nor how generalisable the national estimates are for NSW government schools specifically. It should also be noted that the study reports the prevalence of all possible healthy eating initiatives tested in previous studies, irrespective of the evidence of their effect. Interpretation of the data should consider that the level of implementation of a program does not reflect the effectiveness in improving health outcomes. Additionally, while some survey items were based on valid and reliable tools and all were reviewed by stakeholders, not all measures have been previously validated. Furthermore, the self-report data collection may be a limitation as principals may be too removed from day-to-day classroom practices to answer some survey items accurately (e.g., confectionary used as a classroom reward), as their roles often focus more on administrative and leadership tasks. Self-report data collection may have resulted in social desirability bias, particularly by telephone respondents. Therefore, our findings may potentially overestimate the actual implementation of initiatives assessed. This may warrant further research to validate these data with observational or auditing methods. Finally, data collection occurred in the immediate aftermath of COVID-19. As such, response rates, and implementation of healthy eating initiatives may have been influenced by the COVID-19 burden placed on teaching staff and schools across Australia.^38^ It is difficult to know what the extent of this timing had on our results.

This study has generated evidence to help inform the prioritisation of healthy eating initiatives and to identify evidence-practice gaps to determine where policy or funding support may be warranted. Data from this study allows the level of implementation to be mapped to the evidence base to identify effective healthy eating initiatives not currently implemented at scale. Once evidence-practice gaps are identified we can begin to investigate the barriers to poor implementation and test effective strategies tailored to improve implementation. The findings underscore the need for continued and prioritised support and funding for the implementation of evidence-based primary school healthy eating initiatives not widely implemented.

## 5 Conclusion

The study provides key insights for policymakers and practitioners into the current implementation of various healthy eating initiatives. It provides an understanding of which initiatives are already being implemented across most schools, and insights into which initiatives may benefit from greater policy or resource support. These findings can inform future investments and help identify schools that may need additional support to effectively implement targeted initiatives.

## Data Availability

All data produced in the present study are available upon reasonable request to the authors

## Acknowledgments

The authors would like to thank the participating primary schools and all individuals who contributed to data collection for this study.

## Appendices

## Appendix 1: Research approvals

| Jurisdiction | Approval number |
| --- | --- |
| ACT Department of Education | RES 2314 |
| ACT/NSW Archdiocese of Canberra and Goulburn | NA |
| NSW Department of Education (SERAP) | 2021187 |
| Diocese of Bathurst | NA |
| Diocese of Maitland-Newcastle | NA |
| Diocese of Parramatta | NA |
| Diocese of Wagga Wagga | NA |
| NT Department of Education | 20817 |
| NT Catholic schools | NA |
| QLD Department of Education | 550/27/2550 |
| Archdiocese of Brisbane | 502 |
| Diocese of Cairns | NA |
| Diocese of Rockhampton | NA |
| Diocese of Townsville | 2021-13 |
| SA Department of Education | 2021-0074 |
| SA Catholic schools | 202134 |
| TAS Department of Education | 2022-03 |
| Archdiocese of Hobart | NA |
| VIC Department of Education | 2022_004563 |
| Archdiocese of Melbourne | 1173 |
| Diocese of Ballarat | NA |
| Diocese of Sale | NA |
| Diocese of Sandhurst | NA |
| WA Department of Education | D23/1411981 |
| Catholic Education Western Australia (four Dioceses (Broome, Bunbury, Geraldton, Perth)) | RP2021/44 |

Abbreviations: ACT: Australian Capital Territory; QLD: Queensland; NA: Not Applicable; NSW: New South Wales; NT: Northern Territory; SA: South Australia; SERAP: State Education Research Application Process; TAS: Tasmania; VIC: Victoria; WA: Western Australia

## Appendix 2: Healthy eating initiative definitions

| Opportunities for healthy eating | Recommended strategies | Initiatives | Definitions |
| --- | --- | --- | --- |
| Healthy eating in the classroom | Provide students information they need to choose healthy foods | Healthy eating units of work in PDHPE/HPE curriculum across all year groups | Units of work for teaching healthy eating in the PDHPE/HPE curriculum were included across all year groups |
|  |  | Healthy eating homework provided to students at least once a week to all year groups | Healthy eating homework provided at least once a week to all year groups in the last 12 months |
|  |  | Teachers acted as healthy eating role models for students | Teachers have been acknowledged or role modelled healthy eating in the last 12 months |
|  |  | Students supported to be healthy eating role models for peers | Students have been acknowledged as, or supported to become healthy eating role models in the last 12 months |
|  | Provide opportunities for children to eat healthy foods | Fruit and vegetable breaks in ≥80% of classes daily | Fruit and vegetable breaks are implemented in at least 80% of classes daily |
|  |  | Permission or breaks to drink water in class time in ≥80% of classes daily | At least 80% of classes have permission or breaks to drink water |
|  |  | Ensure confectionary not used as classroom rewards | Confectionary is not used as a reward by teachers in any classes |
|  |  | Implement food growing experiences or food gardens | Food growing experiences or a food garden were provided to students in the last 12 months |
|  |  | Implement healthy cooking programs or similar healthy eating experiences | Healthy cooking or healthy food experiences provided to students in the last 12 months |
|  |  | Healthy eating integrated into at least some key learning areas other than PDHPE | Healthy eating integrated into at least some key learning areas other than PDHPE |
|  |  | Classroom-based programs or strategies on student motivation, goal setting or monitoring of healthy eating | Classroom-based programs and strategies targeting student motivation, goal setting or self-monitoring of healthy eating implemented in the last 12 months. |
| Healthy food available at school | Support and courage children to select healthy foods when purchasing food at school | Provide incentives or rewards to children for purchasing/choosing healthy foods or drinks at school | The school has implemented a reward or incentive system to acknowledge student healthy eating in the last 12 months |
|  |  | School-wide healthy eating programs (e.g. fruit and veg month) | The school has participated in school-wide programs that promoted messages about healthy eating in the last 12 months (e.g. fruit and veg month) |
|  | Ensure foods sold through school food services or events are consistent with dietary guidelines | Energy dense, nutrient poor or sugar-sweetened beverages are not available on the canteen menu | The canteen does not usually sell or provide energy dense, nutrient poor foods or sugar sweetened drinks |
|  |  | Implement healthy fundraising policies | School-wide healthy eating fundraising activities in the last 12 months (e.g. fruit and/or vegetable boxes) |
|  |  | Encourage students to purchase healthier items from canteen (e.g., signalling, positioning, prompting) | At least one strategy is implemented to encourage students to purchase healthier items from the school canteen (either online or in-person) (e.g., colour/traffic light signalling, positioning healthier items more prominently, prompting students to add fruit, vegetables or water to orders, graphical or written feedback to their order) |
|  | Provide healthy foods to children | Provide free breakfast to children | Free breakfast available to all students at least once per week |
|  |  | Provide free fruits and vegetables to children | Free fruits and vegetables available to all students at least once per week to eat at school or take home |
|  |  | Healthy options at school events | Availability of calorie dense, nutrient poor foods and sugar sweetened drinks are at less than half of all school events |
|  |  | Install water stations for free access to cooled plain or optionally carbonated water | Students have access to free cooled water from fountains in the last 12 months |
|  |  | Provision of free healthy school lunches | Free healthy school lunches available to all students over the last 12 months |
|  | Individual counselling | Individual healthy eating advice or counselling for all students (e.g. healthy eating plans) | Individually tailored advice or counselling to improve healthy eating was provided to all students in the last 12 months |
| Healthy food brought to school | Support families to pack healthy lunchboxes | Information and resources to support parents to pack a healthy lunchbox | Healthy lunchbox information was sent home to parents or carers in the last 12 months |
|  | Provide families information about choosing healthy foods | Information on healthy eating sent home to parents (e.g. healthy foods available at the canteen) | Healthy eating information (e.g. healthy lunchboxes, canteens, fruit and vegetable breaks, or school healthy eating policies) sent home to parents or carers in the last 12 months |
|  |  | Workshops with parents on healthy eating | The school conducted workshops for parents/carers which included information on student healthy eating in the last 12 months |
| Healthy eating outside the school or involving families | Support families to purchase and cook healthy foods | Healthy eating programs with an interactive family component | Healthy eating programs (e.g. cooking programs) with an interactive family component delivered to students in the last 12 months |
|  |  | Promote healthy food options at shops close to school | The school has partnered with local stores to promote healthy eating choices to students or parents in the last 12 months |
|  |  | Community members acted as healthy eating role models for students | The school has promoted or engaged a community member as a healthy eating role model in the last 12 months |
|  |  | Healthy eating programs delivered in partnership with community organisations or services | The school partnered with community organisations or services to deliver healthy eating programs in the last 12 months |
| Other |  | The school has a healthy eating plan or policy | The school has a written healthy eating plan or policy |
|  |  | The majority of teaching staff participated in healthy eating professional development | The majority of teaching staff have participated in healthy eating professional development/learning in the last 12 months |
|  |  | Information about school healthy eating strategies/programs shared with school community/education sector | The school has shared information on healthy eating strategies or programs with the school community or sector in the last 12 months |

Abbreviations: PDHPE/HPE: Personal Development, Health and Physical Education

## Appendix C: Principal survey

| Survey item | Responses |
| --- | --- |
| **School and principal characteristics** |  |
| What primary school Year groups does your school cater for? | All = All of the below: Kindergarten/Prep/Reception/Pre-primary year to Year 6<br>K = Kindergarten/Prep/Reception/Pre-primary<br>1 = Year 1<br>2 = Year 2<br>3 = Year 3<br>4 = Year 4<br>5 = Year 5<br>6 = Year 6 |
| Which best describes your current role? | 1 = Executive Principal<br>2 = Principal<br>3 = Deputy Principal<br>4 = Assistant Principal<br>5 = Acting Principal<br>6 = Other role<br>7 = Head of school / Campus / Operations<br>8 = School counsellor / social worker<br>9 = Lead / classroom teacher<br>10 = Health and Wellbeing Teacher / Coordinator<br>11 = Wellbeing and Aboriginal Education Leader |
| What is your current role? |  |
| How long have you been in this role at this school? | 1 = 12 months or more<br>2 = Less than 12 months |
| Please enter the number of years you have been in this role at this school |  |
| Does your school have an operational canteen service? | 1 = Yes<br>2 = No |
| **Healthy eating module** |  |
| For which Year groups does your school's PDHPE/HPE curriculum include programs with units of work for teaching healthy eating? | 1 = Kindergarten/Prep/Reception/Pre-primary<br>2 = Year 1/Year 2<br>3 = Year 3/Year 4<br>4 = Year 5/Year 6<br>All = All Year groups<br>7 = None of the above<br>888 = Unsure |
| For which Year groups does your school have units of work regarding healthy eating in Learning Areas/Key Learning Areas other than PDHPE/HPE (e.g. use of food and nutrition concepts across topic areas)? | 1 = Kindergarten/Prep/Reception/Pre-primary<br>2 = Year 1/Year 2<br>3 = Year 3/Year 4<br>4 = Year 5/Year 6<br>All = All Year groups<br>7 = None of the above<br>888 = Unsure |
| Do classes at your school have specific breaks or have permission to eat vegetables and/or fruit during class time? Note: This could include fruit and vege break, crunch and sip® | 1 = Yes, all classes<br>2 = Yes, some classes<br>3 = No<br>888 = Unsure |
| What percentage of all classes at your school have specific breaks or have permission to eat vegetables and/or fruit during class time? |  |
| On how many days per week does the vegetable and fruit break usually occur in participating classes? | 1 = 1<br>2 = 2<br>3 = 3<br>4 = 4<br>5 = 5<br>6 = Less than 1 day/week<br>7 = None<br>888 = Unsure |
| What percentage of all classes at your school have specific breaks or have permission to drink water during class time? |  |
| How frequently does your school review and update your school canteen menu? | 1 = Every 2 years or less frequently<br>2 = Annually<br>3 = Twice a year (e.g., summer and winter menus)<br>4 = Once a term<br>5 = More than once a term<br>6 = Never - the menu is updated by an external provider<br>888 = Unsure |
| QLD<br>Thinking about your school's current canteen menu, which of the statements are true? | 1 = No drinks with added sugar are available for sale<br>2 = No red items are available on the canteen's everyday menu (red items are foods or drinks low in nutrients and are generally high in energy, saturated fat and/or salt e.g., confectionary, soft drinks, fruit drinks, deep-fried food, iced cakes and slices and doughnuts)<br>3 = The majority of food and drinks available at the canteen are green items (green items are foods or drinks high in nutrients e.g., fruit, vegetables, reduced fat dairy products, lean meat, bread and water)<br>4 = Portion sizes of amber foods and drinks are appropriate for children (amber items are foods and drinks that contain some valuable nutrients but may also be high in energy, saturated fat and/or salt e.g., cereals with added sugar, full fat milk, sweet and savoury snack foods)<br>5 = None of the above<br>888 = Unsure |
| NT<br>Thinking about your school's current canteen menu, which of the statements are true? | 1 = No drinks with added sugar are available for sale<br>2 = No red items are available on the canteen's everyday menu (red items are foods or drinks low in nutrients and are generally high in energy, saturated fat and/or salt e.g., confectionary, soft drinks, fruit drinks, deep-fried food, iced cakes and slices and doughnuts)<br>3 = The majority of food and drinks available at the canteen are green items (green items are foods or drinks high in nutrients e.g., fruit, vegetables, reduced fat dairy products, lean meat, bread and water)<br>4 = Portion sizes of amber foods and drinks are appropriate for children (amber items are foods and drinks that contain some valuable nutrients but may also be high in energy, saturated fat and/or salt e.g., cereals with added sugar, full fat milk, sweet and savoury snack foods)<br>5 = None of the above<br>888 = Unsure |
| NSW<br>Thinking about your school's current canteen menu, which of the statements are true? | 1 = There are no Drinks with added sugar available for sale<br>2 = At least ¾ of the food and drinks available at the canteen are 'everyday' options<br>3 = All available food and drinks meet a health star rating of 3.5<br>4 = Portion sizes of occasional foods and drinks on the canteen menu are appropriate for children<br>5 = None of the above<br>888 = Unsure |
| SA<br>Thinking about your school's current canteen menu, which of the statements are true? | 1 = The majority of food and drinks available at the canteen are green items (green items are foods or drinks high in nutrients e.g., fruit, vegetables, reduced fat dairy products, lean meat, bread and water)<br>2 = Portion sizes of amber foods and drinks are appropriate for children (amber items are foods and drinks that contain some valuable nutrients but may also be high |
|  | in energy, saturated fat and/or salt e.g., cereals with added sugar, full fat milk, sweet and savoury snack foods)<br>3 = No sugar sweetened, or artificially sweetened drinks (except milk) are sold<br>4 = No red items are available for everyday purchase<br>5 = None of the above<br>888 = Unsure |
| VIC<br>Thinking about your school's current canteen menu, which of the statements are true? | 1 = No drinks with added sugar are available for sale<br>2 = No red items are available on the canteen's everyday menu (red items are foods or drinks low in nutrients and are generally high in energy, saturated fat and/or salt e.g., confectionary, soft drinks, fruit drinks, deep-fried food, iced cakes and slices and doughnuts)<br>3 = The majority of food and drinks available at the canteen are green items (green items are foods or drinks high in nutrients e.g., fruit, vegetables, reduced fat dairy products, lean meat, bread and water)<br>4 = Portion sizes of amber foods and drinks are appropriate for children (amber items are foods and drinks that contain some valuable nutrients but may also be high in energy, saturated fat and/or salt e.g., cereals with added sugar, full fat milk, sweet and savoury snack foods)<br>5 = None of the above<br>888 = Unsure |
| TAS<br>Thinking about your school's current canteen menu, which of the statements are true? | 1 = No drinks with added sugar are available for sale<br>2 = No red items are available on the canteen's everyday menu (red items are foods or drinks low in nutrients and are generally high in energy, saturated fat and/or salt e.g., confectionary, soft drinks, fruit drinks, deep-fried food, iced cakes and slices and doughnuts)<br>3 = The majority of food and drinks available at the canteen are green items (green items are foods or drinks high in nutrients e.g., fruit, vegetables, reduced fat dairy products, lean meat, bread and water)<br>4 = Portion sizes of amber foods and drinks are appropriate for children (amber items are foods and drinks that contain some valuable nutrients but may also be high in energy, saturated fat and/or salt e.g., cereals with added sugar, full fat milk, sweet and savoury snack foods)<br>5 = None of the above<br>888 = Unsure |
| ACT<br>Thinking about your school's current canteen menu, which of the statements are true? | 1 = No drinks with added sugar are available for sale<br>2 = No red items are available on the canteen's everyday menu (red items are foods or drinks low in nutrients and are generally high in energy, saturated fat and/or salt e.g., confectionary, soft drinks, fruit drinks, deep-fried food, iced cakes and slices and doughnuts)<br>3 = The majority of food and drinks available at the canteen are green items (green items are foods or drinks high in nutrients e.g., fruit, vegetables, reduced fat dairy products, lean meat, bread and water)<br>4 = Portion sizes of amber foods and drinks are appropriate for children (amber items are foods and drinks that contain some valuable nutrients but may also be high in energy, saturated fat and/or salt e.g., cereals with added sugar, full fat milk, sweet and savoury snack foods)<br>5 = None of the above<br>888 = Unsure |
| WA<br>Thinking about your school's current canteen menu, which of the statements are true? | 1 = No drinks with added sugar are available for sale<br>2 = No red items are available on the canteen's everyday menu (red items are foods or drinks low in nutrients and |
|  | are generally high in energy, saturated fat and/or salt e.g., confectionary, soft drinks, fruit drinks, deep-fried food, iced cakes and slices and doughnuts)<br>3 = 60% or more of the food and drinks available at the canteen are green items (green items are foods or drinks high in nutrients e.g., fruit, vegetables, reduced fat dairy products, lean meat, bread and water)<br>4 = Portion sizes of amber foods and drinks are appropriate for children (amber items are foods and drinks that contain some valuable nutrients but may also be high in energy, saturated fat and/or salt e.g., cereals with added sugar, full fat milk, sweet and savoury snack foods)<br>5 = None of the above<br>888 = Unsure |
| Does your school usually sell or provide any of the following in the school canteen? | 1 = Fruit<br>2 = Vegetables i.e., vegetable sticks, (not salad on a sandwich)<br>3 = Water<br>4 = Regular soft drinks i.e., not diet<br>5 = Diet soft drinks<br>6 = Less than 99% fruit juice<br>7 = 100% fruit juice<br>8 = Reduced fat milk drinks<br>9 = Flavoured milks<br>10 = Energy drinks<br>11 = Other sugar sweetened drinks (e.g., cordials, flavoured mineral waters, sports drinks, iced teas, sweetened waters, sports waters, flavoured crushed ice drink)<br>12 = Confectionary e.g., lollies, chocolate<br>13 = Sweet or savoury biscuits/cakes/muffins/donuts or muesli bars<br>14 = Potato crisps or other salty snacks such as Twisties or corn chips<br>15 = Deep fried foods<br>16 = Ice-creams covered in chocolate<br>17 = Other ice-creams, ice blocks or frozen desserts<br>18 = Savoury pastries i.e., pies and sausage rolls<br>19 = None of the above<br>888 = Unsure |
| What proportion of food and drinks actively marketed at your school canteen are green items? Note: Active marketing includes product placement, specials and lowering prices to increase sales, and promotion through printed and digital media* | 1 = All<br>2 = Most (more than half but not all)<br>3 = Some (but less than half)<br>4 = None<br>888 = Unsure |
| How many of the occasional food and drinks your canteen advertises would you estimate are healthy? Note: Including via meal deals, specials, commercial branding, at point of sale, or at eye level of shelves and counters† | 1 = All<br>2 = Most (more than half but not all)<br>3 = Some (but less than half)<br>4 = None<br>888 = Unsure |
| How many of your school canteen/tuckshop staff (paid or volunteer) have participated in training, or accessed support services (e.g., Canteen Helpline, Menu Check Services) in providing healthy food and drink options at school in the last 12 months? | 1 = All staff<br>2 = Most staff (more than half but not all)<br>3 = Some staff (but less than half)<br>4 = None<br>888 = Unsure |
| Has your school participated in or provided students with any of the following opportunities in the last 12 months? | 1 = Healthy fundraising activities (e.g., vegetable boxes)<br>2 = Food growing experiences or a food garden<br>3 = Healthy cooking programs or similar healthy food experiences for students<br>4 = Participated in a healthy eating campaign, such as National Nutrition Week or Fruit and Veg Month |
|  | 5 = Free healthy school lunches available to all students<br>6 = Regularly provided students free sugar-free beverages (other than water)<br>7 = Students have free access to water fountains with cooled water<br>8 = None of the above<br>888 = Unsure |
| How often have sugar sweetened drinks been available to purchase or provided to students at school events such as discos, sporting carnivals and camps in the last 12 months? Note: Sugar sweetened drinks include cordials, sports drinks, sweetened waters | 1 = All school events<br>2 = Most school events (more than half but not all)<br>3 = Some school events (but less than half)<br>4 = None<br>888 = Unsure |
| How often have red items been available or provided to students at school events such as discos, sporting carnivals and camps in the last 12 months?† | 1 = All school events<br>2 = Most school events (more than half but not all)<br>3 = Some school events (but less than half)<br>4 = No classes<br>888 = Unsure |
| In what proportion of classes do teachers at your school provide confectionary as rewards? | 1 = All classes<br>2 = Most classes (more than half but not all)<br>3 = Some classes (but less than half)<br>4 = No classes<br>888 = Unsure |
| Does your school provide free fruit and vegetables to students to eat at school or take home at least once a week? | 1 = Yes, available to all students every day<br>2 = Yes, available to all students at least once a week<br>3 = Yes, but only available to selected students every day<br>4 = Yes, but only available to selected students at least once a week<br>5 = Sometimes, but not regularly<br>6 = No, never<br>888 = Unsure |
| Does your school provide free breakfast to students? Note: This could include via school canteen or external programs/organisation such as Samaritans, Foodbank SA, KickStart | 1 = Yes, available to all students every day<br>2 = Yes, available to all students at least once a week<br>3 = Yes, but only available to selected students every day<br>4 = Yes, but only available to selected students at least once a week<br>5 = Sometimes, but not regularly<br>6 = No, never<br>888 = Unsure |
| Has your school partnered with any local stores to promote their available healthy eating choices to students or parents in the last 12 months? Note: This could include via newsletters | 1 = Yes<br>2 = No<br>888 = Unsure |
| Has your school implemented any healthy eating programs or strategies for students which include an interactive parent/family/carers component in the last 12 months? Note: This could include parent/child workshops or cooking classes that involve active participation | 1 = Yes, interactive cooking program(s)<br>2 = Yes, healthy eating<br>3 = No<br>888 = Unsure |
| Please enter the program name(s) or a brief description of the strategies with interactive components. Note: Please separate program name(s)/strategies with a comma |  |
| Has your school implemented any reward or incentive systems to acknowledge student healthy eating, in the last 12 months? Note: Small rewards or incentives could include sports equipment, water bottles, a merit system | 1 = Yes<br>2 = No<br>888 = Unsure |
| Please describe any reward or incentive systems that acknowledged student healthy eating in the last 12 months |  |
| Do you currently use any of the following strategies to encourage students to purchase healthier items from the canteen? | 1 = Adding coloured symbols to indicate healthier and less healthy menu options (Menu labelling)<br>2 = Arranging menu items to make healthier options more prominent (Positioning) |
|  | 3 = Encouraging users to add fruit, vegetables or water to their orders (Prompting)<br>4 = Graphical or written feedback to show users how healthy their order is (Feedback)<br>5 = Other strategy<br>6 = None<br>7 = Price signalling<br>8 = promotion through school communication or posters<br>888 = Unsure |
| What other canteen strategies does your canteen use to encourage healthy purchases? |  |
| Do you currently use any of the following strategies to encourage students to purchase healthier items from the <u>online canteen</u>? | 1 = Adding coloured symbols to indicate healthier and less healthy menu options (Menu labelling)<br>2 = Arranging menu items to make healthier options more prominent (Positioning)<br>3 = Encouraging users to add fruit, vegetables or water to their orders (Prompting)<br>4 = Graphical or written feedback to show users how healthy their order is (Feedback)<br>5 = Other strategy<br>6 = None<br>7 = Price signalling<br>8 = promotion through school communication or posters<br>888 = Unsure |
| What other online canteen strategies does your online canteen use to encourage healthy purchases? |  |
| Are there any other programs or strategies that your school implements targeting healthy eating that we haven't asked you specifically about? | 1 = Yes<br>2 = No<br>888 = Unsure |
| Please describe the other programs or strategies that your school implements which targets healthy eating |  |
| **Healthy eating and physical activity module** |  |
| For which Year groups has your school provided students with weekly healthy eating and/or physical activity homework in the last 12 months? | 1 = Kindergarten/Prep/Reception/Pre-primary – healthy eating<br>2 = Kindergarten/Prep/Reception/Pre-primary – physical activity<br>3 = Year 1/Year 2 - healthy eating<br>4 = Year 1/Year 2 - physical activity<br>5 = Year 3/Year 4 - healthy eating<br>6 = Year 3/Year 4 - physical activity<br>7 = Year 5/Year 6 - healthy eating<br>8 = Year 5/Year 6 - physical activity<br>9 = None of the above<br>888 = Unsure |
| Has your school sent information home to parents/carers about any of the following healthy eating and/or physical activity topics in the last 12 months? Note: This could include via letter, newsletter, pamphlet, email, website or parent meetings | 1 = Healthy lunchboxes<br>2 = Healthy canteen<br>3 = Vegetable, fruit and water breaks<br>4 = Benefits of, or strategies to increase physical activity<br>5 = School policy or guidelines regarding healthy eating<br>6 = Limiting small screen recreation<br>7 = Benefits of safe active travel<br>8 = Encouragement to 'park and walk' with their children to school<br>9 = School policy or guideline regarding physical activity<br>10 = Other<br>11 = None of the above<br>12 = Benefits of, or strategies to increase healthy eating or water consumption<br>888 = Unsure |
| What other healthy eating and/or physical activity topics has your school sent information home to parents/carers about? |  |
| Has your school conducted any workshops for parents/carers (either face-to-face or online) which included information on student healthy eating and/or physical activity in the last 12 months? | 1 = Yes, safe walking, cycling, scootering or other active travel activities<br>2 = Yes, strategies to improve student healthy eating<br>3 = Yes, strategies to improve student physical activity<br>4 = Yes, other<br>5 = No, did not conduct any workshops<br>6 = yes, strategies to improve general wellbeing including healthy eating and physical activity<br>888 = Unsure |
| Please describe the other workshops for parents/carers |  |
| What proportion of your teaching staff have participated in healthy eating professional development/learning in the last 12 months? | 1 = All staff<br>2 = Most staff (more than half but not all)<br>3 = Some staff (but less than half)<br>4 = No staff<br>888 = Unsure |
| Does your school have a policy written by the school (i.e., not the Education sector or state-wide policy) promoting healthy eating and nutrition? Note: This can include a general school policy that explicitly references healthy eating and nutrition practices | 1 = Yes<br>2 = No<br>888 = Unsure |
| Has your school shared information on your healthy eating and/or physical activity strategies or programs with your school community or education sector in the last 12 months? Note: School community or education sector could include a School Board, P&C, or school education director/principal/region director/executive group | 1 = Yes, healthy eating<br>2 = Yes, physical activity<br>3 = No, neither<br>888 = Unsure |
| Has your school provided students with individually tailored counselling or advice to increase their healthy eating and/or physical activity in the last 12 months? Note: This could include online programs, activity plans developed with teachers | 1 = Yes, healthy eating to all students<br>2 = Yes, healthy eating to selected students<br>3 = Yes, physical activity to all students<br>4 = Yes, physical activity to selected students<br>5 = None of the above<br>888 = Unsure |
| Has your school participated in any school-wide initiatives that promoted messages about healthy eating and/or physical activity in the last 12 months? (e.g., Fruit and Vege month, Jump Rope for Heart). Note: Ways of promoting messages school-wide could include putting up posters around school, hosting school events - this does not include strategies that only included providing information to parents | 1 = Yes, healthy eating<br>2 = Yes, physical activity<br>3 = No, neither<br>888 = Unsure |
| Have any students at your school been acknowledged as, or supported to become healthy eating and/or physical activity role models in the last 12 months? | 1 = Yes, healthy eating<br>2 = Yes, physical activity<br>3 = No, neither<br>888 = Unsure |
| Have any staff members at your school been acknowledged as, or acted as healthy eating and/or physical activity role models in the last 12 months? | 1 = Yes, healthy eating<br>2 = Yes, physical activity<br>3 = No, neither<br>888 = Unsure |
| Has your school promoted or engaged any people outside of your school community as healthy eating and/or physical activity role models in the last 12 months? (e.g., local or well-known athlete) | 1 = Yes, healthy eating<br>2 = Yes, physical activity<br>3 = No, neither<br>888 = Unsure |
| Has your school partnered with any community organisations or services to deliver programs targeting student healthy eating and/or physical activity in the last 12 months? Note: Community organisations or services could include working with local sports clubs or local community consultation to develop programs | 1 = Yes, healthy eating<br>2 = Yes, physical activity<br>3 = No, neither<br>888 = Unsure |
| Has your school implemented any classroom-based programs or strategies targeting student's motivation, personal goals or self-monitoring of healthy eating and/or physical activity in the last 12 months? Note: Classroom-based programs or strategies could include activity plans, journals either in class or at home | 1 = Yes, healthy eating<br>2 = Yes, physical activity<br>3 = No, neither<br>888 = Unsure |
| Are there any other programs or strategies that your school implements targeting healthy eating or physical activity that we haven't asked you specifically about? | 1 = Yes<br>2 = No<br>888 = Unsure |
| Please describe the other programs or strategies that your school implements which targets healthy eating or physical activity |  |

Abbreviations: ACT: Australian Capital Territory; QLD: Queensland; NA: Not Applicable; NSW: New South Wales; NT: Northern Territory; PDHPE/HPE: Personal Development, Health and Physical Education; PE: Physical Education; SA: South Australia; TAS: Tasmania; VIC: Victoria; WA: Western Australia
\*Participants in all states and territories were asked this survey item, except those in NSW
†Only participants in NSW were asked this survey item

## Appendix 4: Summary of out-of-scope schools

| Reason for being out of scope | n |
| --- | --- |
| Taking part in other trial | 210 |
| Not invited due to research approval not being extended (NSW only) | 846 |
| School closed / no current staff or students | 5 |
| Distance schools | 2 |
| Does not accept phone calls | 1 |

Abbreviations: NSW: New South Wales

## Appendix 5: Associations between implementation of healthy eating initiatives and school characteristics

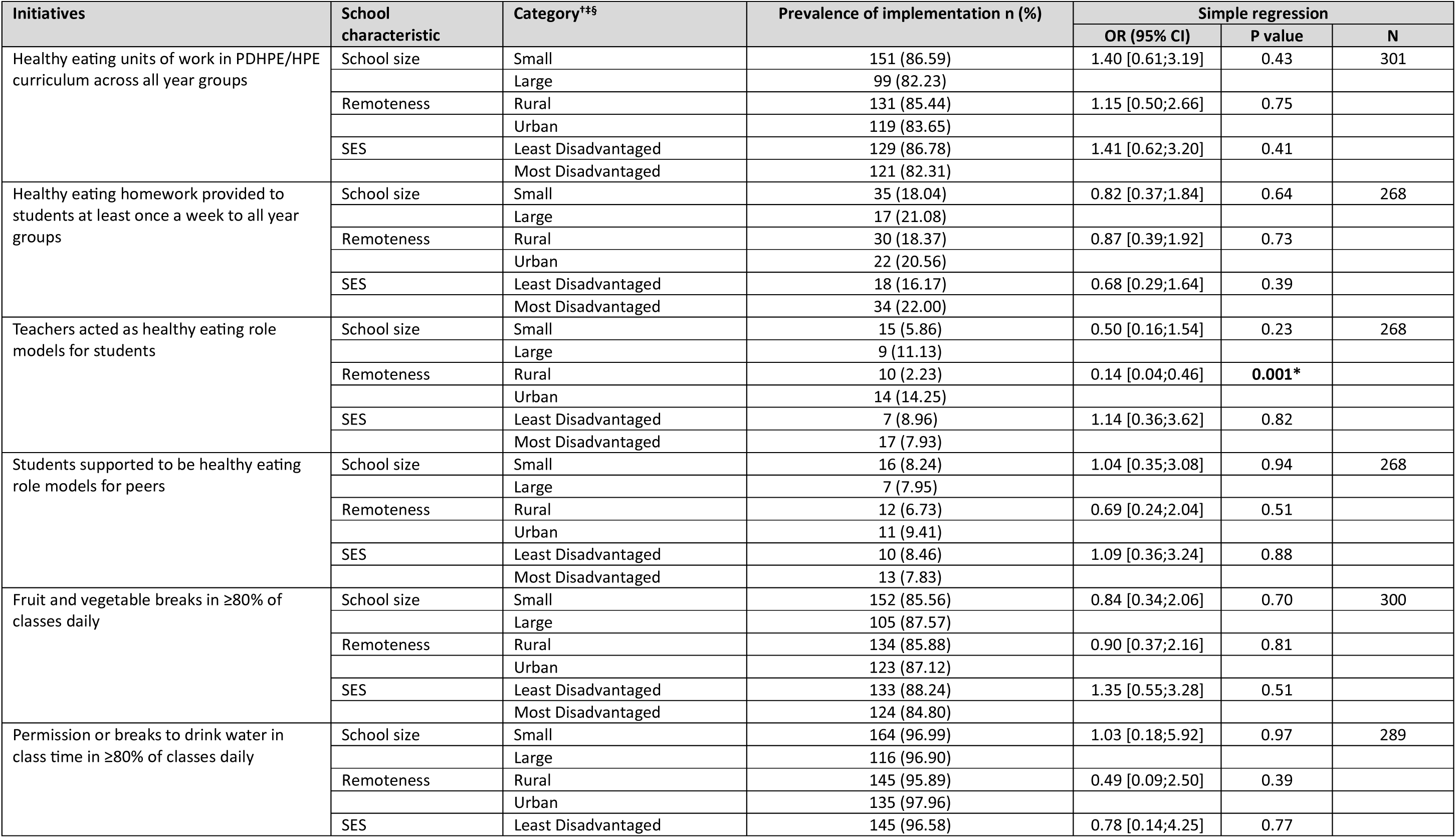
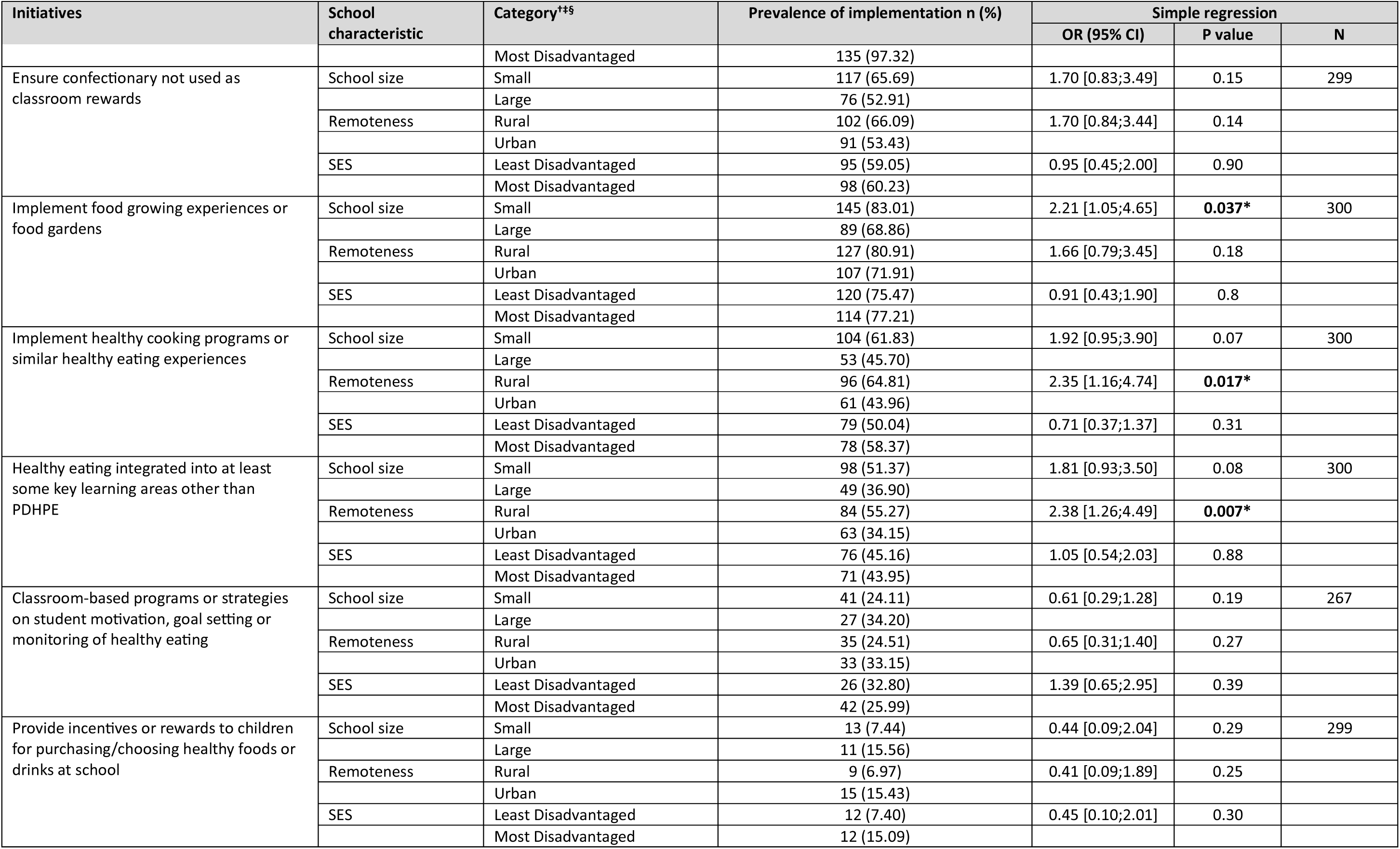
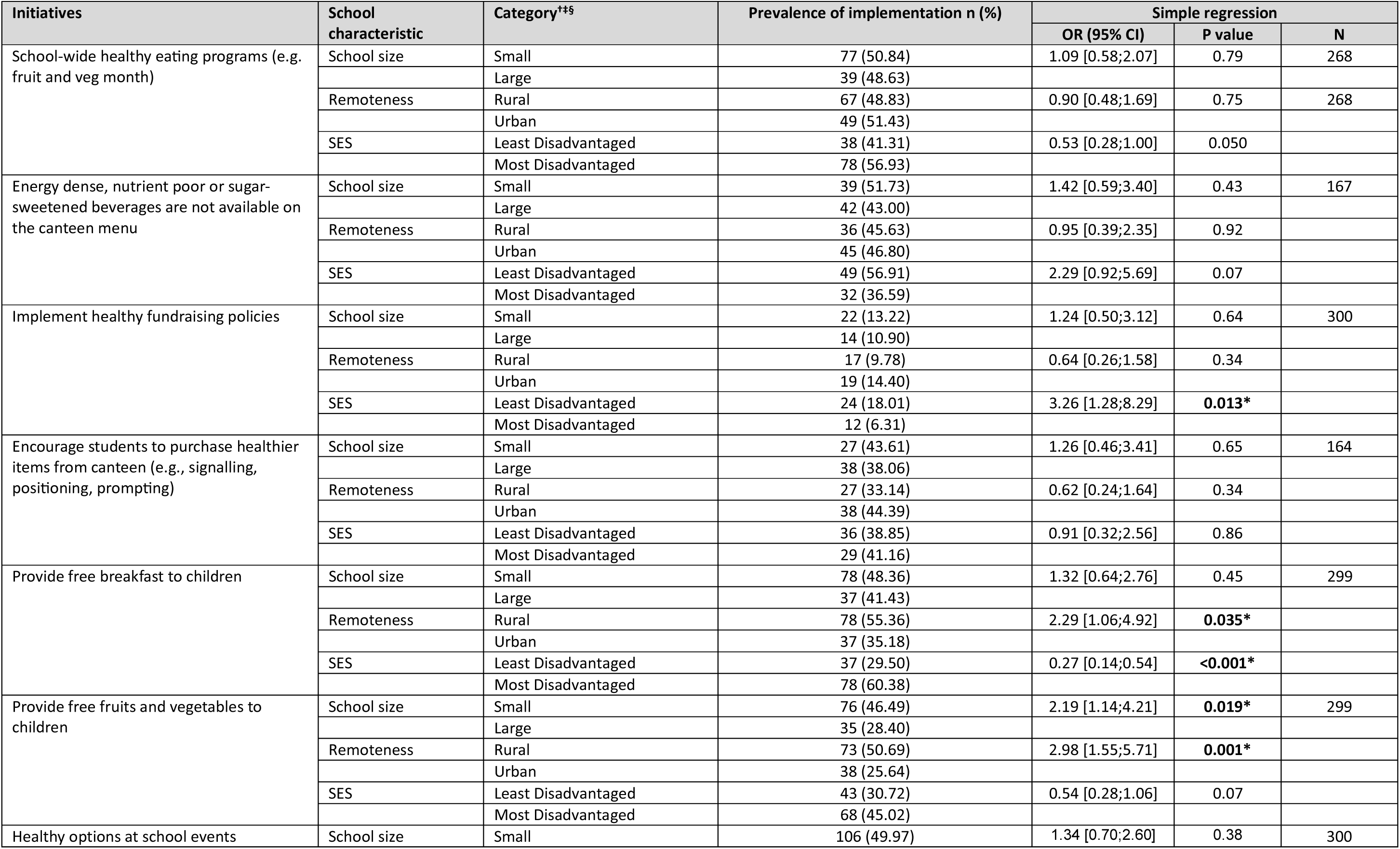
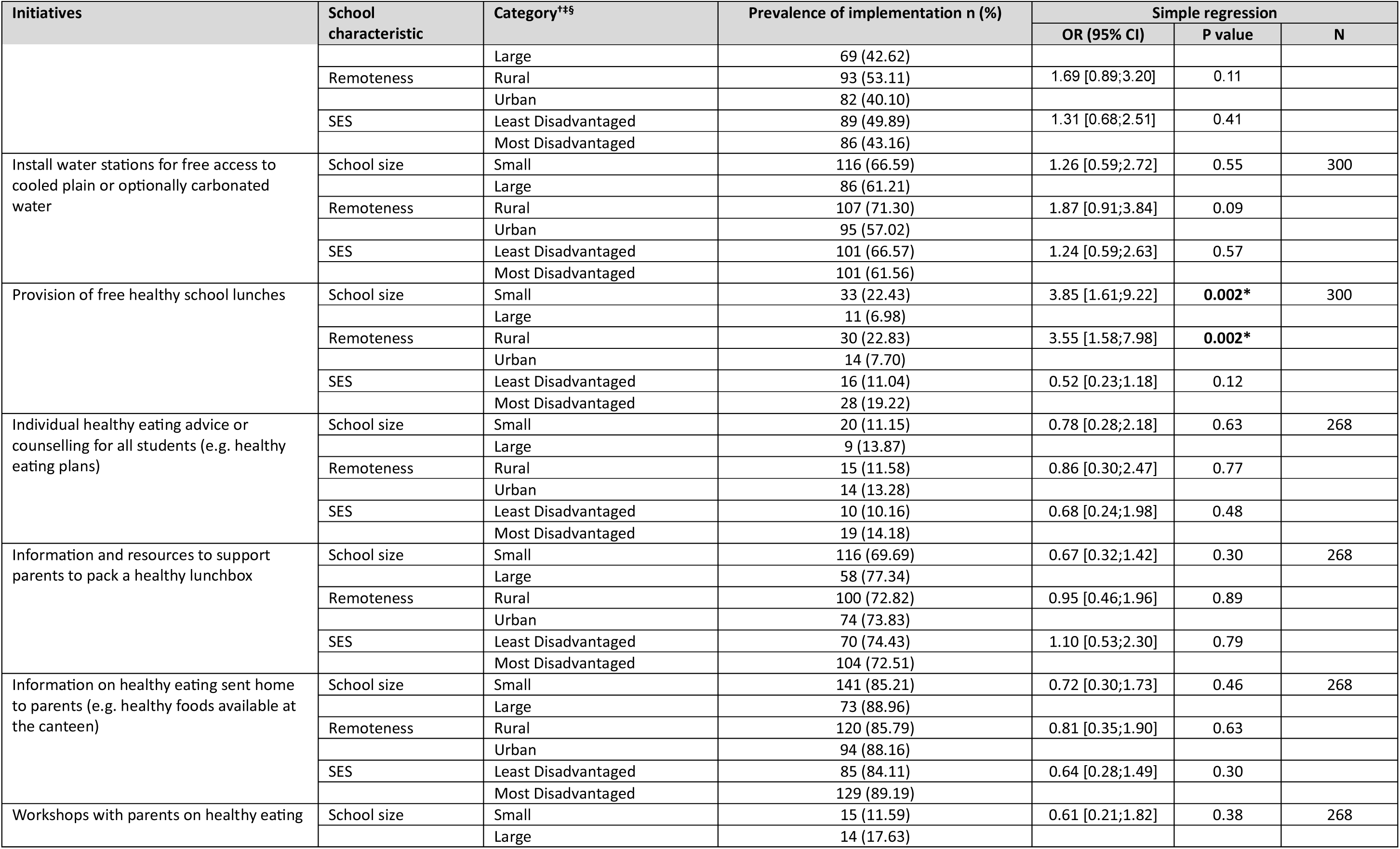
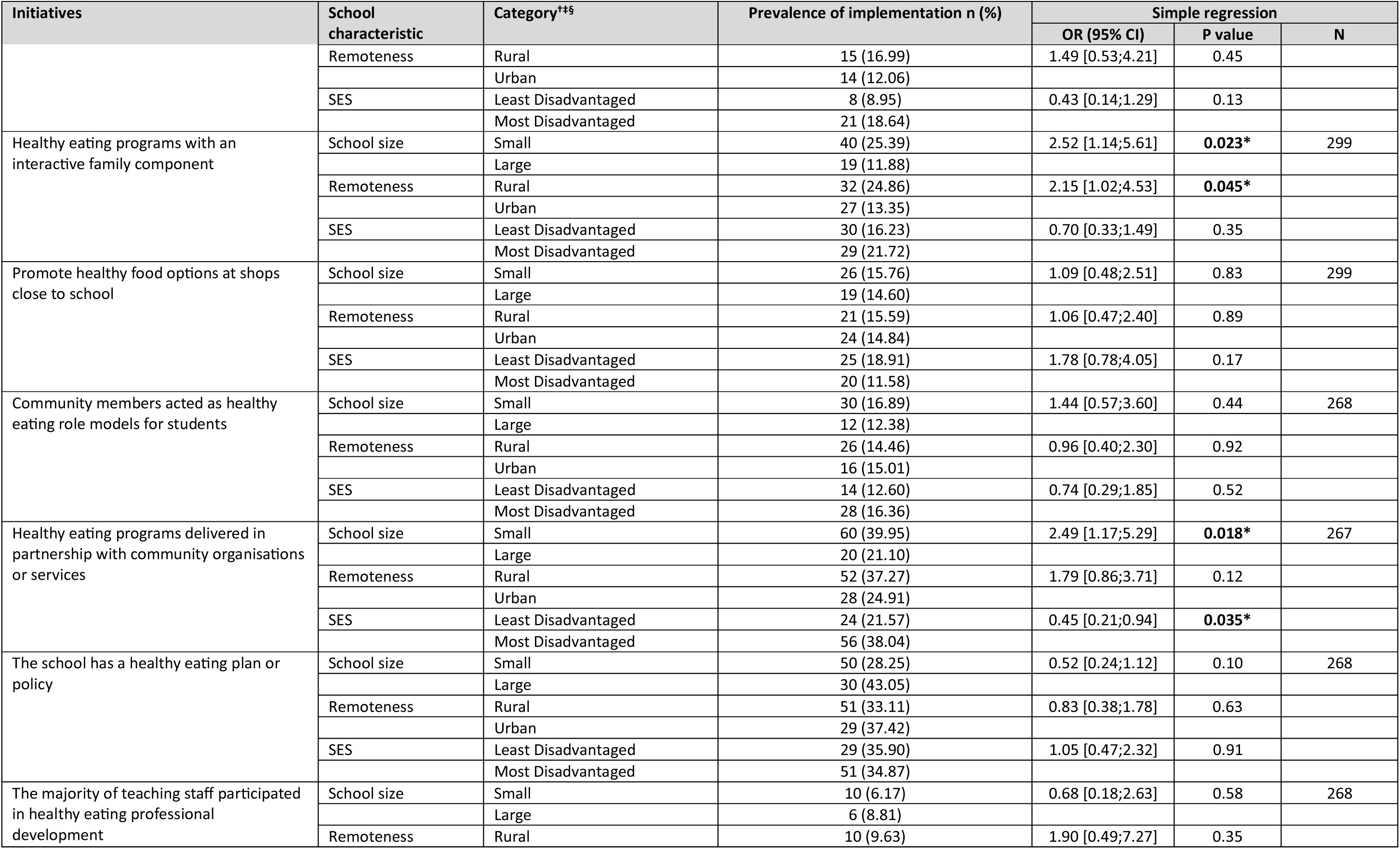
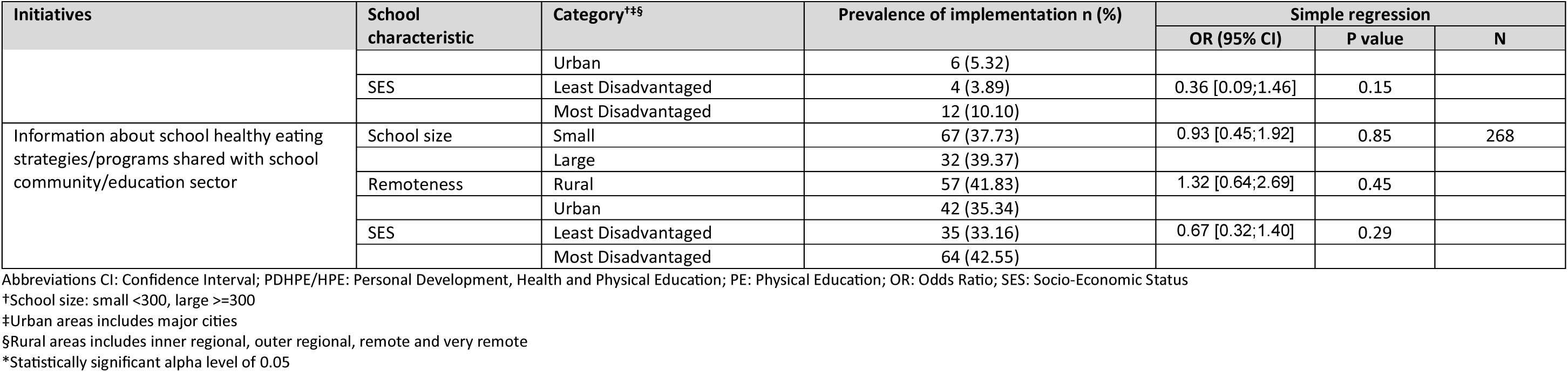

